# Monogenetic rare diseases in biomedical databases and text mining

**DOI:** 10.1101/2022.04.07.22273575

**Authors:** Anastasia Nesterova, Eugene Klimov, Sergey Sozin, Vladimir Sobolev, Peter Linsley, Pavel Golovatenko-Abramov

## Abstract

The testing of pharmacological hypotheses becomes faster and more accurate, but at the same time more difficult than even two decades ago. It takes more time to collect and analyse disease mechanisms and experimental facts in various specialized resources. We discuss a new approach to aggregating individual pieces of information about a single disease using Elsevier’s automated text mining technology. Developed algorithm allows for the collection of published facts in a unified format starting only with the name of the disease. The special template, which combines research and clinical descriptions of diseases was developed. The approach was tested, and information was collected for 55 rare monogenic diseases. Clinical, molecular, and pharmacological characteristics of diseases with supporting references from the literature are available in the form of tables and files. Manually curated templates for 10 rare diseases, including top ranked Cystic Fibrosis and Huntington’s disease, were published to demonstrate the results of the described approach.

## 2 Introduction

Diseases, even monogenic ones, have complex cellular and clinical mechanisms described in hundreds of published experimental papers, case-studies, and population studies. Successful biomedical research requires access to large amounts of information. Diagnosis and treatment of many diseases are improving thanks to large volumes of results from genomics, proteomics, pharmacology, and systems biology data available online. However, the search and analysis of new discoveries has become a complex and long-term task.

Published collections and books give only a superficial view of the state of research, as they usually do not systematically cover the results of experimental studies or clinical trials and do not discuss new scientific hypotheses. Alternatively, accumulated information about human diseases is available in many public and commercial databases, which can be divided broadly into 2 categories: databases with medical or with biological information. The first category includes classifications and taxonomies of diseases, symptoms, phenotypes, supported by international associations (ICD11 – International Classification of Diseases, 11^th^ Revision, https://icd.who.int/); scientific institutes (MESH – Medical Subject Headings 2021, https://meshb.nlm.nih.gov/, DO – Disease Ontology, https://disease-ontology.org/); or private companies (SNOMED, MedDra). Collections of clinical trials (https://clinicaltrials.gov) and decisions of regulatory organizations (FDA) also belong to the first category. Second category includes many directories, on-line applications, and databases, public and commercial. For example, collections about links between gene mutations and diseases (OMIM, TCGA); pathological changes in gene expression in cells and tissues (HPA); drug targets and biomarkers (KEGG – Kyoto Encyclopaedia of Genes and Genomes, http://www.kegg.jp/); and many others. Several reviews of such databases can be found in the literature (Santos et al., 2017; Wang et al., 2019; Zaman et al., 2017).

In a special category we include resources that collect and combined information from other sources. These can be the resources that aggregate information from other databases (for example, MalaCards, https://www.malacards.org/); resources specialized on a disease or type of diseases (for example, Orphanet, https://www.orpha.net/); databases that store information from individual publications (Resnet – 2020, https://www.pathwaystudio.com). Unfortunately, many resources cease due to loss of funding or support after a few years or months of accumulating information.

There are several common problems across all categories of sources about human diseases. The search within the databases can be inefficient and inaccurate due to mistakes during the complex task of correctly combining terms and data. The lack of references to specific publications and experiments in databases often calls into question the validity of the data presented in them. Finally, it is difficult to find a resource that properly combines several different types of facts about the disease – medical, pharmacological, clinical, and molecular. These problems force a doctor, a researcher, or a student to check the information in many sources and then spend time on cleaning the data. Not surprisingly, many medical researchers prefer to use proven methods – reading reviews and monographs.

In this work, we describe our approach of automating the collection of facts about rare monogenic diseases in clinical, molecular, and pharmacological areas to create templated disease descriptions with links to original sources and published references.

Both monogenic and rare disease are an artificial concept, the definition of which depends on medical schools and traditions (Chong et al., 2015; Venugopal et al., 2018). Monogenic diseases are usually determined by the type of heredity such as autosomal or sex-linked dominant types (with two copies of mutant genes) or recessive types (with one copy) (Chial, 2008). Rare diseases are defined by their prevalence. There is no generally accepted understanding in the world of what number should determine the rarity of the disease, since the statistical calculations depend on the size of populations, countries, and nationalities. However, the prevalence of a rare disease is 3-6 percent or one to six people per 10,000 people (Nguengang Wakap et al., 2020). European legislation defining a prevalence of rare disease as a threshold of not more than 5 affected persons per 10,000 (“Procedural document: Epidemiology of rare diseases in Orphanet,” 2019). However in the USA, disease may be considered as a rare with incidence of less than 200,000 patients, in Brazil with 65 cases per 100,000, in Russia with no more than 10 cases per 100,000 people (Hedley, 2018).

Orphanet, which is developed by European organizations, is the biggest resource about rare diseases (Weinreich et al., 2008) and includes more 6 000 disease names. In USA-based biggest databases which link individual genes with disease phenotypes (OMIM and ClinVar), there are 7,315 monogenic rare disease reported. It is worth noting that, despite significant progress, for the majority of rare diseases with Mendelian heredity, a link with genetic causes has not yet been found (Chong et al., 2015).

In summary, available data about rare and genetic diseases require complex expert analysis and interpretation to allow separating quality data from speculative ones. Methods of how to summarize and visualize facts and make them accessible to a wider range of specialists are now more relevant than ever.

## 3 Results and discussion

### 3.1 Ranking of rare monogenic diseases

By monogenic disease we understand those phenotypes for which it is known that one or a limited number of mutations cause and determine the development of the disease. We used publicly available epidemiologic data from the Orphanet database to define the list of monogenic diseases. Orphanet’s publicly available file (http://www.orphadata.org) includes 6043 unique diseases. For 1245 of them (including chromosome level and microdeletions diseases) are not genetically inherited or no inheritance data available. 156 diseases are marked as disease with multiple inheritance pattern. In total, we consider 3330 diseases as monogenic, which were marked with label of “autosomal” or “sex-linked” and are not “multigenic”. Some diseases in Orphanet have several contradicting inheritance labels (for example, autosomal dominant and not genetically inherited). We count all known inheritances regardless other contradiction labels. To have the ranked list of rare monogenetic diseases by their social importance, we count number of data points for the disease in Orphanet by several criteria multiplied by priority weight (k):

- disease labeled rare “worldwide” (k=5)
- disease has known point prevalence (k=1)
- disease prevalence is several patients on 10 or 100 thousand people (k=3)
- disease data validated (k=4)
- counts of evidence for the disease in Orphanet (k=1)
- disease marked with all priority prevalence criteria (worldwide; with point prevalence 1-6 persons in 10 or 100 thousand) (k=6).

Point prevalence measured worldwide was our priority criteria for ranking monogenic diseases. Point prevalence is a quantity of cases scaled up to the general population at a given time. Other measurements of prevalence are annual incidence (number of newly diagnosed cases in a population in one year) and birth prevalence (number of cases observed at birth relative to the number of children born alive at a given moment). Point prevalence is assigned to the higher number of diseases in Orphanet and, also, it is considered as the valid measure of a disease being or not being rare in human population (Nguengang Wakap et al., 2020) (Figure 1).

**Figure 1.**
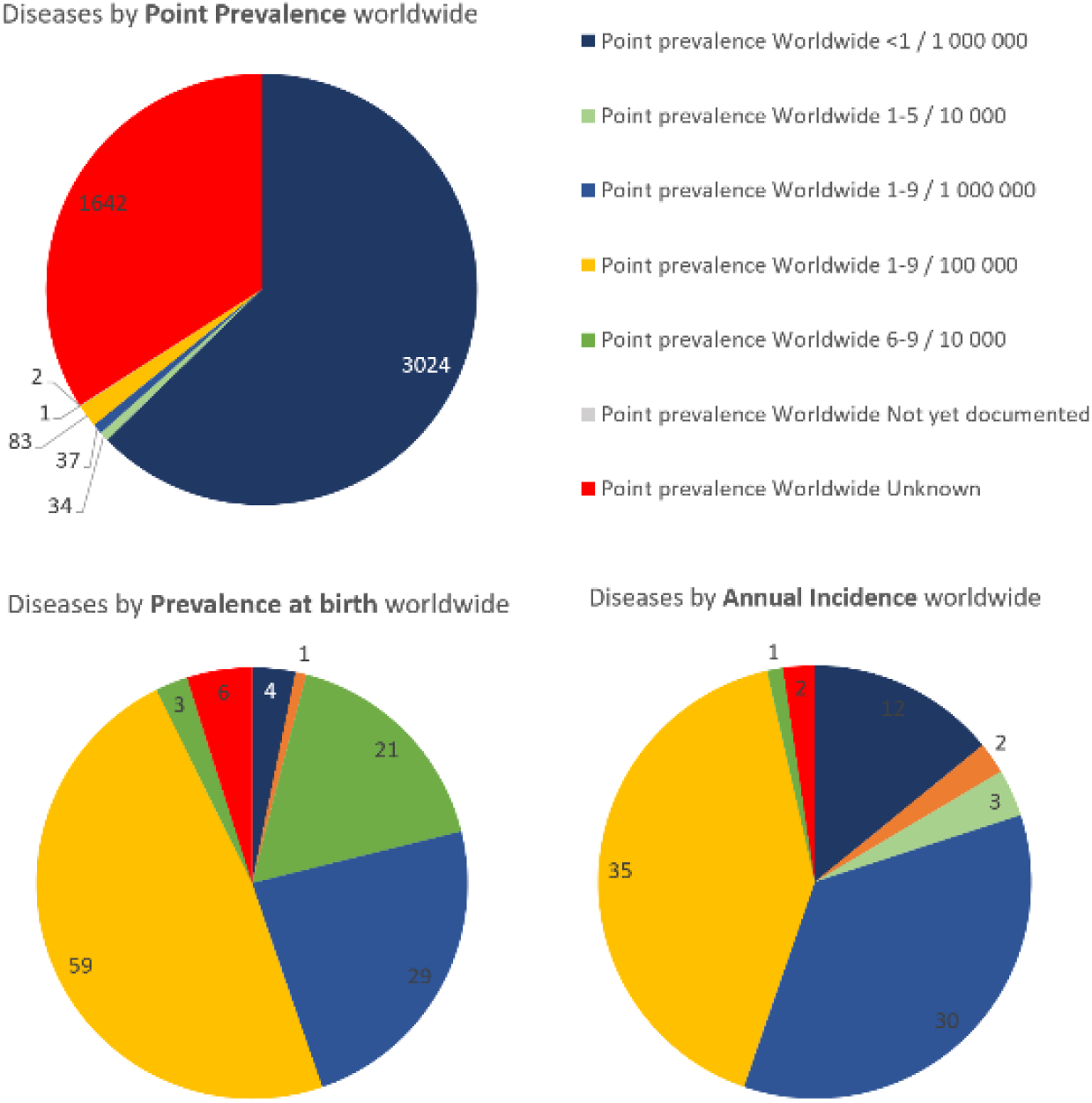
Distribution of rare monogenic diseases by types of prevalence in Orphanet.

By the prevalence ranking approach in the scale from 1 to 100, only 23 diseases had ranks higher than 10. Only 45 monogenic diseases met all priority criteria and pointed to diseases which are not extremely rare and not very common worldwide (See supplemental materials, file “Rare Monogenic diseases”). Hemophilia A was the top ranked disease by prevalence score mostly due to the greatest number of country-specific evidence in Orphanet.

For the next step, rare monogenic diseases were ranked by literature score, i.e., how well the disease is studied and presented in the literature. For this we used Elsevier text mining tools and Resnet 2020 network. Final ranking combined prevalence and literature scores and scaled them in 1 to 100 points. Only 33 diseases had highest scores which are the 1^st^ percentile of all data points (Figure 2). Cystic Fibrosis, hemophilia, retinitis pigmentosa had highest combined scores, i.e. they are the most social important rare monogenic diseases according to our ranking method. A complete list of diseases ranked by prevalence and text-analysis available in Supplemental materials.

**Figure 2.**
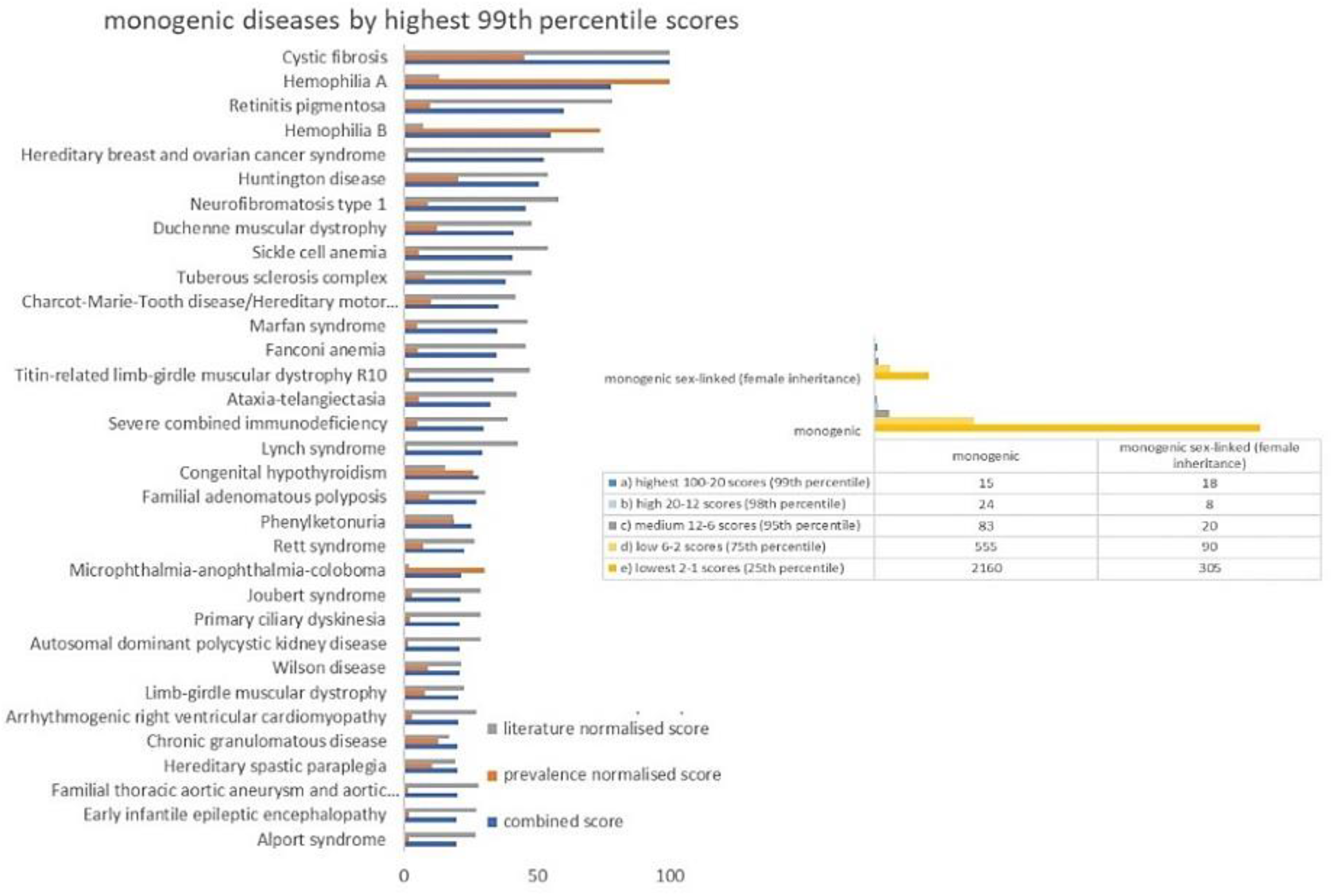
Top ranked monogenic rare diseases by combination of point prevalence score and literature coverage.

In different sources, the names and classifications of diseases may be different. Therefore, at the beginning of the work, we checked how names of monogenic diseases are represented in different sources. Public disease identifiers are the most convenient and reliable way to compare data. We used the OrphanetIDs to find matching diseases in the Elsevier literature database Resnet and in Elsevier Text Mining (ETM) tools. Public identifiers from OMIM, Malacards, MESH, ICD11, Disease Ontology are also widely used for matching medical terms. Pharmapendium (PP, https://www.pharmapendium.com/) did not contain disease identifiers, and diseases were matched by their names (see Methods). As a result, we found that all top ranked diseases from Ophanet are represented in Resnet taxonomy and can be used for ETM. Only a small proportion of rare monogenic diseases were represented in PP and clinicaltrials.gov. Based on the analysis and ratings, 20 rare monogenic diseases were selected for the next stage of work, designing the templates (Table 1).

**Table 1.** Top scored monogenic rare diseases selected for curated templates (see Supplemental file “Monogenic diseases” for more details).

| orhanet id | rare monogenic disease | combined score | point prevalence | connectivity (facts in literature database) |
| --- | --- | --- | --- | --- |
| 586 | Cystic fibrosis | 100.0 | 1-9 / 100 000 | 4573 |
| 98878 | Hemophilia A | 77.78865 | 1-9 / 100 000 | 573 |
| 791 | Retinitis pigmentosa | 60.3 | 1-5 / 10 000 | 3571 |
| 145 | Hereditary breast and ovarian cancer syndrome | 52.6457 | 1-5 / 10 000 | 3432 |
| 399 | Huntington disease | 50.84916 | 1-9 / 100 000 | 2446 |
| 442 | Congenital hypothyroidism | 28.18924 | 1-5 / 10 000 | 664 |
| 716 | Phenylketonuria | 25.5261 | 1-9 / 100 000 | 815 |
| 778 | Rett syndrome | 22.91543 | 1-9 / 100 000 | 1180 |
| 905 | Wilson disease | 20.72394 | 1-9 / 100 000 | 945 |
| 342 | Familial Mediterranean fever | 13.12671 | 1-5 / 10 000 | 657 |
| 886 | Usher syndrome | 12.5756 | 1-9 / 100 000 | 284 |
| 15 | Achondroplasia | 10.91765 | 1-9 / 100 000 | 302 |
| 269 | Facioscapulohumeral dystrophy | 10.32049 | 1-9 / 100 000 | 408 |
| 906 | Wiskott-Aldrich syndrome | 8.688134 | 1-9 / 1 000 000 | 370 |

### 3.2 MOLECULAR AND CLINICAL TEMPLATE OF DISEASE DESCRIPTION

Disease templates are files with facts, text, tables, and images in highly structured and user-friendly formats which save researchers and authors time in creating or writing new content about a given disease for research or clinical use and deepening the value of this content with accessible data and reference links, all available in one source. Example uses of these templates include using them as a starting point to write a book chapter on a rare disease topic, or to write a disease overview for a web page. We developed a special template format which is a combination of research and clinical disease description. Templating allowed us to create and share well-referenced content on rare disease quickly and effectively.

Six general categories were chosen to summarize facts from different sources about the disease and combine clinical, pharmacological, and molecular descriptions (Table 2). The categories were chosen based on the disease description scheme presented in public resources and based on possibilities for an automation. Instead of using clinical descriptions written by experts, we included lists and rankings of key symptoms and parameters, as well as text mining results.

**Table 2.** Molecular and Clinical Disease Template. Abbreviations: ORH (Orphaned database), PS (Pathway Studio Resnet-2020 database), ICD11 (International Classification of Diseases, 11th database), ETM (Elsevier Text Mining tool), CK (Clinical Key database), PP (PharmaPendium database), CT (ClinicalTrial database)

| Section | Sub-section | Sources |
| --- | --- | --- |
| **1. Terminology** | a) Definition | ORH, MedlinePlus |
|  | b) Synonyms | PS |
|  | c) Codes | ICD11, MESH, ORH, OMIM, CUI, MalaCards |
|  | d) Classification | DO, PS |
| **2. Epidemiology/ Demographics** | a) Prevalence | ETM |
|  | b) Risk Factors | ETM, CK |
| **3. Clinical Presentation/ Diagnosis** | a) Early symptoms | ETM |
|  | b) Other symptoms | ETM, CK |
|  | b) Complications | ETM, CK |
|  | c) Differential Diagnosis | ORH, ETM, CK |
|  | d) Diagnostic Procedure | ORH, ETM, CK |
| **4. Etiology/ Pathology** | a) Causes | ORH, MedlinePlus, ETM |
|  | b) Genetics | PS, ETM |
|  | c) Biomarkers | PS |
|  | d) Pathways | PS |
| **5. Treatment/ Follow Up** | a) Non-pharmacological Therapy | ETM |
|  | b) Pharmacological Therapy | PP, ETM |
|  | c) Drug Repurposing | PS |
|  | d) Ongoing Research | CT, PS |
|  | e) Prognosis | ETM, CK |
|  | f) Routine Monitoring | ETM, CK |
| **6. Case Studies** | a) Case Studies | ETM, PubMed |

### 3.3 DATA COLLECTION AND ANALYSIS FOR MONOGENIC DISEASES

Monogenetic rare diseases are poorly represented in most public resources or in replicated information from Orphanet, one of the most complete databases for rare diseases. For disease templates we used Orphanet as a starting point and then added the results of text mining of published papers and information from Elsevier resources (publicly available), which were not present in non-commercial databases.

Automatic text mining reduces time and effort and gets maximum results from published medical texts, case-reports, experimental data, patents, and systematic reviews. There are many technologies for automatic text reading that may achieve high quality of extracting facts or citations from articles in a simplified and structured way (Mishra et al., 2014; Wang et al., 2021).

For filling disease templates with sentences and citations about characteristics of diseases (molecular, clinical, pharmacological, etc.), we used ETM (https://demo.elseviertextmining.com), which was built around MedScan technology (Nesterova et al., 2020; Novichkova et al., 2003). MedScan text mining has high confidence scores (recall and precision) for annotating biomedical facts and retrieving quates from the literature (F1 scores of 90.6 for disease concepts, 87.4 for anatomy concepts, 91.1 in general for combined molecular, medical, pharmacological concepts) (not published internal evaluation).

The search was limited to the first 20 quotes in the last 5 years (in case of insufficient number of results, filters by number and date were removed). API of Elsevier Text Mining and Advanced Query Language (AQL) were used to automate the search (see Methods). For all disease template sections except Symptoms and Complication, we used the advanced query type “relation(…)” – which search for specific semantic links between two or more terms (for example, disease name and symptoms). For Symptoms and Complication sections, the queries type “sentence(…)” and “paragraph(…)” which search terms anywhere in the sentence and anywhere in the paragraph, respectively, gave better results than relation query type. The queries about diagnostic procedure and non-pharmacological treatments retrieved the most information on average. The less informative queries were the ones about the symptoms and causes of the disease (Table 3).

**Table 3.** Performance of text mining queries about rare diseases.

|  | **Template Subsection** | **Query Sensitivity/ Specificity\*** | **Average\***<br>**\*** |
| --- | --- | --- | --- |
| 1 | Epidemiology/Demographics/Prevalence (incidence) | low/broad | 4.7 |
| 2 | Epidemiology/Demographics/Risk factors | high/broad | 13.7 |
| 3 | Clinical presentation/Diagnosis /Symptoms early | high/narrow | 9.0 |
| 4 | Clinical presentation/Diagnosis/Symptoms others common most | high/broad | 1.8 |
| 5 | Clinical presentation/Diagnosis / Complications | high/broad | 11.2 |
| 6 | Clinical presentation/Diagnosis/Differential Diagnosis | high/broad | 13.4 |
| 7 | Clinical presentation/Diagnosis/Diagnostic procedure | high/broad | 22.1 |
| 8 | Etiology /Pathology /Causes | high/broad | 7.8 |
| 9 | Treatment/Follow Up/Other treatments (non-pharmacological) | low/broad | 20.5 |
| 10 | Treatment/Follow Up/Prognosis | high/narrow | 13.1 |
| 12 | Case studies | high/narrow | 15.7 |

*\*Sensitivity shows the proportion of non-relevant results is retrieved. Queries with narrow specificity are focuses on retrieval of one meaning of the term; with broad specificity - several different meanings and includes related terms. \*\*Average of supporting sentences by one disease (48 diseases total).*

The way a query was composed, and the choice of keywords influenced the result; however, in our opinion, the popularity of the research topic over the past 5 years was the main reason that some queries retrieved more information. For example, in demographic studies, hereditary breast and ovarian cancer syndrome had the highest number of sentences. Also, congenital hypothyroidism which is an umbrella term for several syndromes had the highest number of sentences which discussed causes of the disease (Figure 3). Supporting references for rare diseases retrieved using Elsevier Text Mining were published and can be download in Mendeley Data (see Supplemental files for links).References for sub-sections can be extracted automatically with ETM API for any disease name with the code available in GitHub (see Supplemental materials).

**Figure 3.**
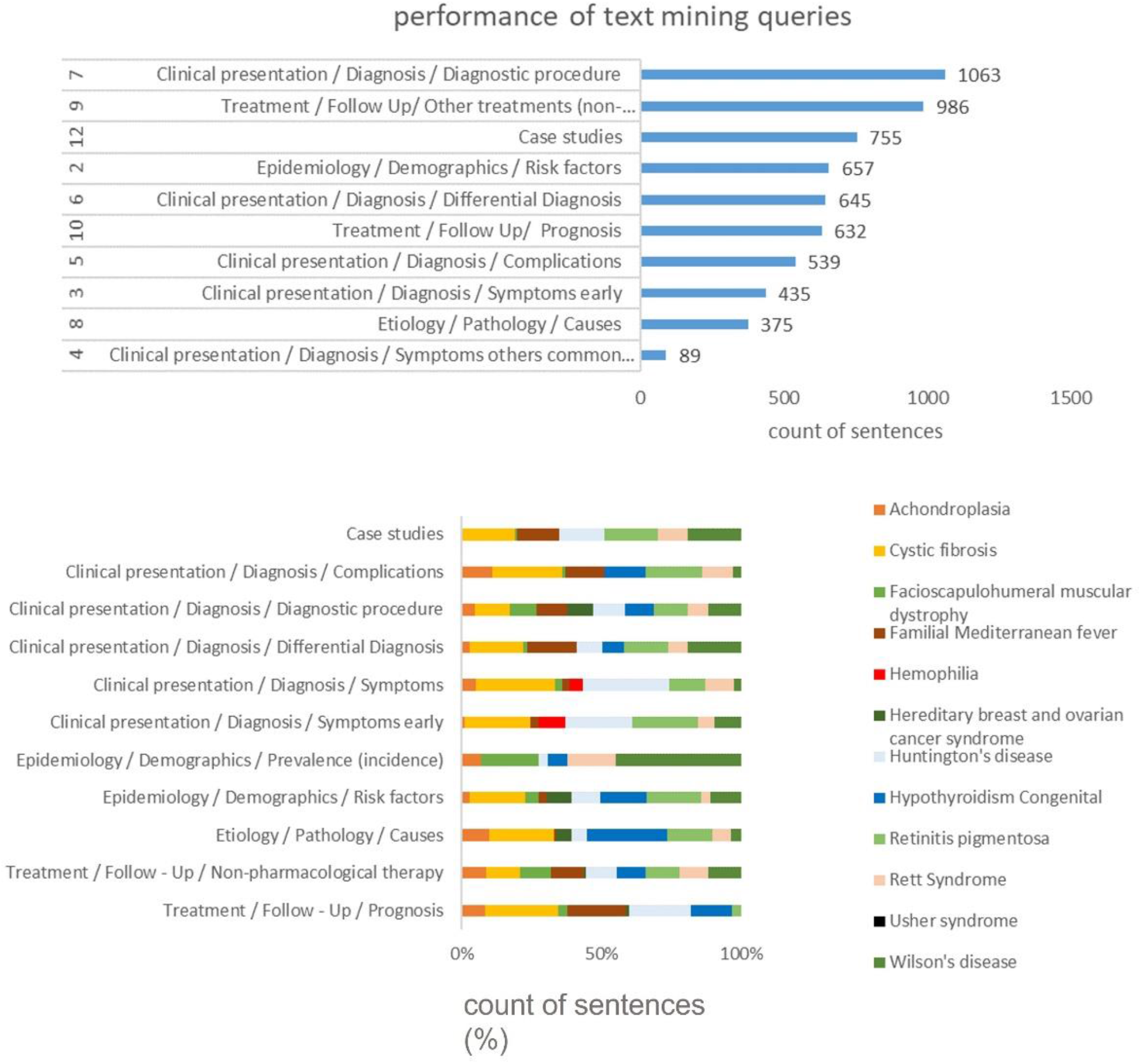
Performance of Elsevier Text Mining API which retrieved published in literature information about 48 rare diseases with.

By definition, monogenic rare diseases are supposed to have fewer of initial gene mutations or molecular alterations that drive the development of the symptoms. Etiology/Pathology section includes lists of mutated genes, known biomarkers and cell signaling (pathways) which may describe molecular causes of the disease. We used commercial software Pathways Studio (https://www.pathwaystudio.com/) and biomedical network Resnet – 2020 (Nesterova et al., 2020) which contains associations and links between proteins, genes, diseases and other concepts extracted from the literature based on the descried above text mining technology (Figure 4). Higher connectivity number in the network (Table 1) indicates that the disease has more known associated proteins, cell processes, drugs, clinical parameters, or anatomy concepts.

**Figure 4.**
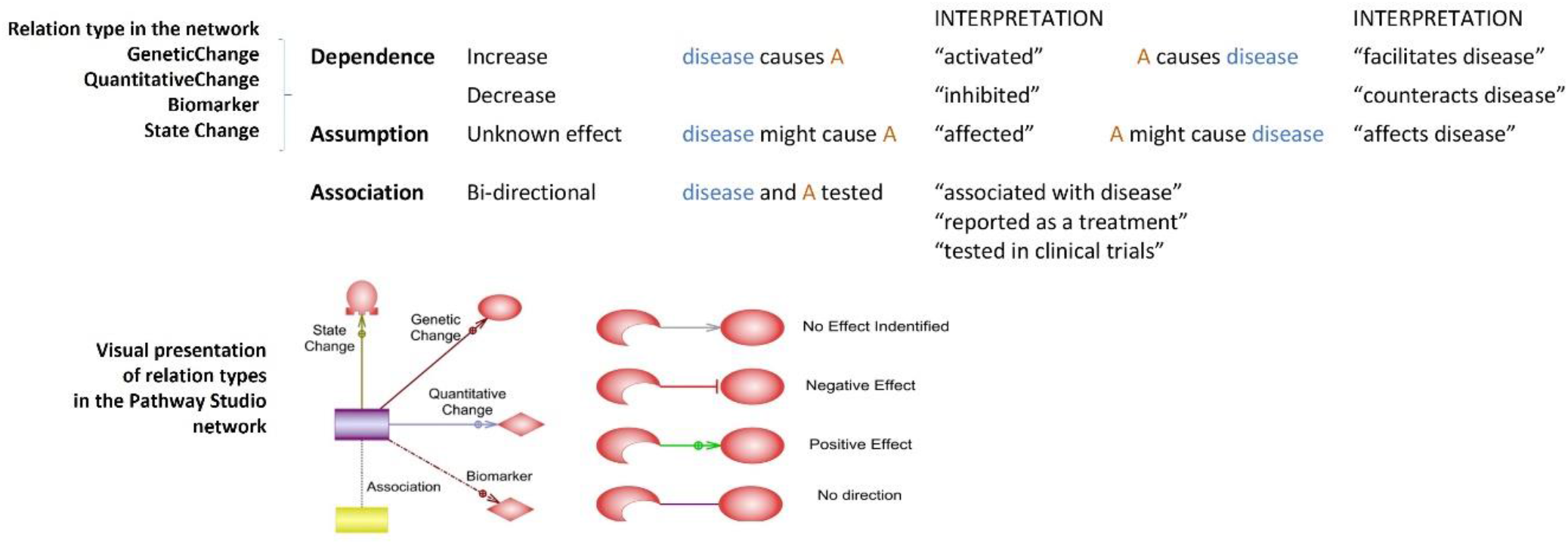
Types of relationships between disease and molecules in Resnet – 2020 literature network. “A” is an entity in the network and can be a mutated gene, protein, biomarker, cell, tissue, organ, drug or small molecule, cell process.

Resnet network keeps not only the facts of association between two concepts but also direction and effect of their relationships. This allowed us to assign and categorise mutated genes and biomarkers by their functional effects on the disease progression (“associated with disease”, “facilitates disease”, “activated” or “inhibited in disease”, “affects disease”, and in rare cases “counteracts disease” (Figure 4).

From the network search results, we included in templates top 10 genes with mutations and 10 single nucleotide variants (SNVs) sorted by number of supporting sentences (see Methods). For the next step, we searched for molecular and anatomy biomarkers that describe proteins, cell, tissues, organs which may indicate the progression of the diseases or linked with their symptoms, and which are typically defined in papers as “biomarkers”. Biomarkers were sorted by number of sentences, known effect of relation, and most recent year of publication.

Finally, we included in disease templates pathways and sub-network that were curated manually and can be used for visualization and as sources of references for each reported association between the disease and genes for example. The result of Resnet-2020 search for selected rare diseases can be downloaded as tables or in Biological Pathway Taxonomy (See Supplemental materials).

Pharmacological data in templates include list of known FDA-approved drugs as well as drugs in research from text mining and clinical studies. Top 10 drugs were selected for templates sorted by number of references in each relation in Resnet-2020. We assigned 4 categories for drugs: “reported as a treatment”, “tested in completed clinical trials”, and “tested in recruiting clinical trials”.

To enrich disease templates with drug-centered information, we also used published data from Pharmapendium database which contains information about drugs and active ingredients approved in Europe and in the US. For disease templates, we used publicly available information about toxicity and adverse effects with dose and route of administration; and about known drug targets or cell systems tested for the drug. Number of clinical studies and endpoints tested for drug and the disease, as well as number of recruiting clinical trials and completed clinical trials were exported from the ClinicalTrials (clinicaltrial.gov).

### 3.4 HUNTINGTON’S DISEASE EXAMPLE

Cystic Fibrosis and Huntington’s diseases had highest scores (100 and 50) in the list of social significant rare monogenic diseases. Diseases had high literature scores (100 and 54) thus were well presented in used sources.

“Huntington’s disease” is a broad term that typically describes classical Huntington’s disease. There are other rare forms of the disease including juvenile Huntington disease, late onset Huntington disease, akinetic-rigid form of Huntington’s disease. According to National Library of Medicine, Huntington disease is a progressive brain disorder that causes uncontrolled movements, emotional problems, and loss of thinking ability (cognition) (https://medlineplus.gov). Basal ganglia disease, extrapyramidal disorder, disorder presenting primarily with chorea are parent taxonomical categories for the disease.

The second Epidemiology/Demographics section of the Huntington’s disease template included citations to the prevalence which is defined as 3 to 7 per 100,000 people of European ancestry by NCBI or as 1 patient to 20 000 – 100 000 persons by Orphanet. In addition, in 2020 meta-analysis defined incidence rate of HD of 0.648 per 100 000 patient-years (Sienes Bailo et al., 2020).References about clinical presentation of the Huntington’s disease selected for the template indicate mental problems for symptoms and complications and molecular diagnostics and brain physiological test for diagnostics (Table 4).

**Table 4.** Major clinical citations from the Huntington’s disease template

| Section | Evidence | Source |
| --- | --- | --- |
| Complications | Depression, Loss of Ability to Interact with Others, At risk of harming others, self-mutilation, impaired self-care | Clinical Key (sorted by prevalence) |
| Diagnostic procedure | neurologic examination, MRI of brain, Molecular Diagnostic Testing, magnetic resonance imaging, history and physical examination, CT scan, positron emission tomography (PET) brain study, CT of brain | Clinical Key (sorted by prevalence) |
| Risk factors | "Three major categories of risk factors for the onset and progression of HD were CAG repeat length, CAG instability, and genetic modifiers." | ETM, (Krewski et al., 2017) |
|  | "Genetic factors were determined to be the dominant risk factors for HD, with only weak associations for environmental or demographic factors reported from a limited number of studies." | (Chao et al., 2017) |
| Symptoms | "Emotional dysregulation (irritability, anger/aggression, and anxiety) and increased inflammation are early emerging symptoms which can be detected decades before the onset of motor symptoms in HD patients." | (Raper et al., 2016) |

**Table 5.** Pathways from Elsevier Pathway Collection about Huntington’s disease (see Supplemental materials)

| **Name** | **Description** | **Entitie**<br>**s** | **Relation**<br>**s** |
| --- | --- | --- | --- |
| Huntingtin Gene Mutation in Striatal Neuron | Huntington's disease is a progressive, fatal, neurodegenerative disorder caused by an expanded CAG repeat in the huntingtin gene (HGT). | 48 | 80 |
| Axonal Transport Impairment in Huntington Disease | Normal huntingtin (HTT) was shown to play a role in fast axonal transport. | 33 | 50 |
| NF-kB Signaling Activation in Huntington Disease | IKBKB is a prominent regulator of neuroinflammation, which is implicated in the pathogenesis of Huntington disease. | 20 | 28 |
| PPARGC1A Repression in Huntington Disease | Mutant huntingtin causes transcriptional repression of the mitochondrial PPARGC1A. | 30 | 52 |
| Glutamate Toxicity Effects on Striatum Medium Spiny Neurons in Huntington's Disease | Huntington's disease is a neurodegenerative disorder, striatum is one of the most affected regions. Glutamate and dopamine signalling pathways induce Ca<sup>2+</sup> overload and apoptosis of neurons. | 63 | 78 |
| ER Stress in Huntington Disease | Misfolded huntingtin (HTT) induces endoplasmic reticulum (ER) stress contributing to pathogenesis of Huntington Disease. | 27 | 39 |
| Huntingtin Mutant Protein Toxicity Effects | Mutant HTT (mHTT) is cleaved at several points to generate toxic fragments with abnormal compact alpha conformation. Proteolytic cleavage of mHTT may be one of the keys for disease pathogenesis. | 16 | 22 |

**Table 6.** Top substances from published studies for Huntington’s disease treatments.

| **Top drugs or substances reported for the treatment** | **Most recent PMID** | **Most recent year** | **Most recent sentence from publications** |
| --- | --- | --- | --- |
| **Minocycline** | 30873213 | 2019 | "Furthermore, there is rapidly growing evidence suggesting that minocycline may have some neuroprotective activity in various experimental models such as cerebral ischemia, traumatic brain injury, amyotrophic lateral sclerosis, Parkinson's disease (PD), Huntington's disease, and multiple sclerosis." |
| **Tetrabenazine** | 33197487 | 2020 | "Tetrabenazine and Olanzapine are often prescribed drugs to treat Huntington's disease but victims are in dire need of a permanent treatment of this terrific disorder." |
| **Resveratrol** | 29542037 | 2019 | "Overall, resveratrol attenuates the symptoms of Parkinson's, Huntington's and Alzheimer's disease in rodents." |
| **ACR-16** | 33164941 | 2020 | "Pridopidine is an investigational drug for Huntington disease." |
| **laquinimod** | 32270922 | 2020 | "Laquinimod has been found to have therapeutic effects on multiple sclerosis and Huntington's disease." |
| **sertraline** | 26511378 | 2015 | "These findings with our current study suggest that the sertraline may protect neurons by increase BDNF and/or 5-HT in Huntington's disease, though we could not differentiate whether sertraline's protection is directly mediated by 5-HT or indirectly by increasing BDNF levels." |
| **4-phenyl-butyrate** | 23711791 | 2015 | "4-phenyl butyric acid is a FDA approved drug for treatment of urea cycle disorders and is under investigation in pre-clinical and clinical models for treatment of Alzheimer disease, cystic fibrosis, Huntington disease, cancer, spinal muscular atrophy and additional motor neuron disorders." |
| **Dimebon** | 23027934 | 2012 | "Second, Dimebon has been the subject of extensive clinical studies in both Alzheimer's disease and Huntington disease." |
| **riluzole** | 30526230 | 2019 | "Interestingly, riluzole stimulated BDNF release from human platelets (Türk and Frizzo, 2015) and increased BDNF production in patients with Huntington disease (Squitieri et al., 2009)". |

The fourth section combines evidence on the pathology of the disease. Genetic causes are known for Huntington’s disease, and its molecular mechanisms are well-studied. Mutations in HTT (Huntingtin) gene (expansion of guanine-adenine-guanine (GAG) repeats) is a known major cause of Huntington’s disease (Tell-Marti et al., 2017). However other genes and proteins were also reported to be involved in disease mechanisms (Figure 5).

**Figure 5.**
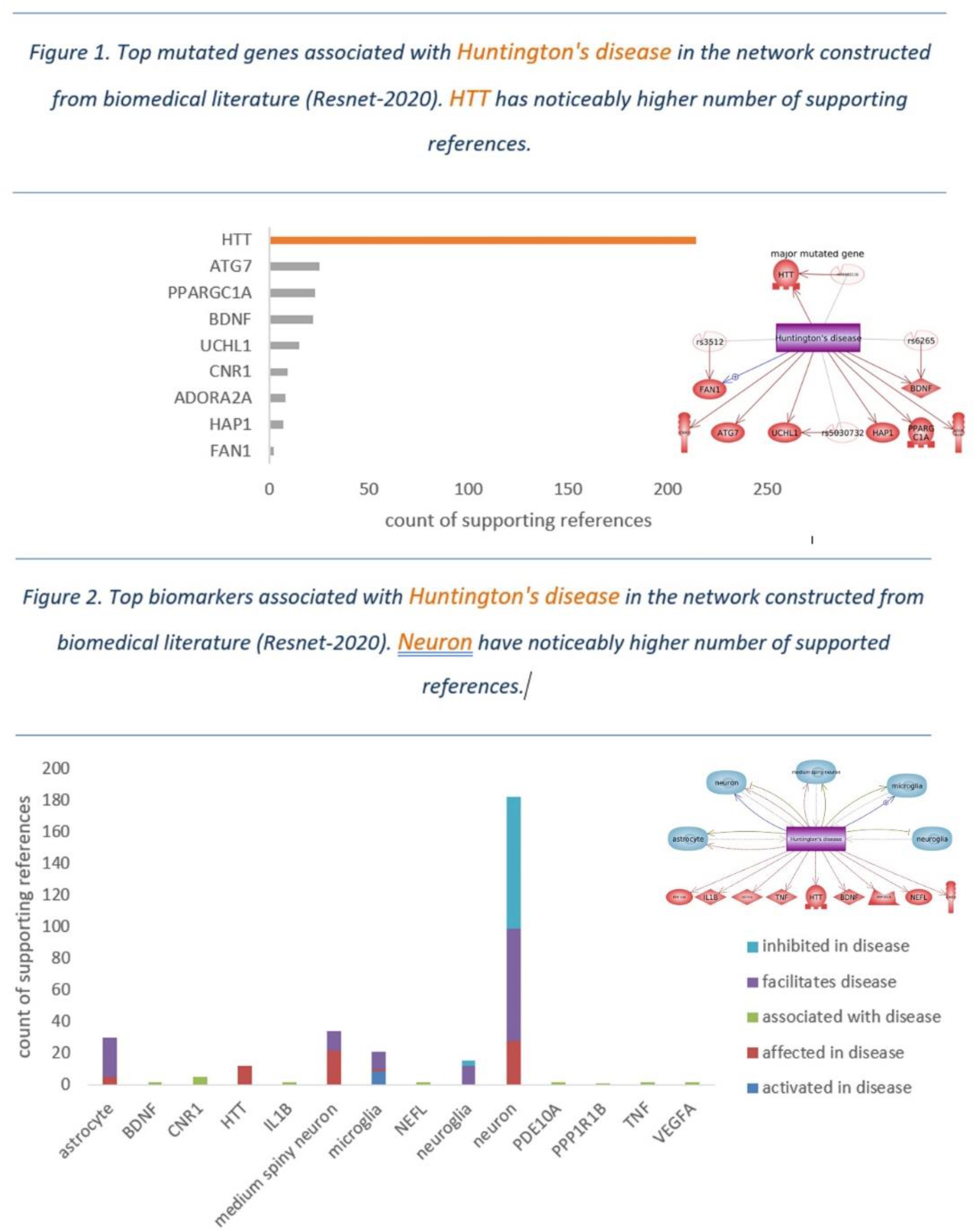
Infographics from Huntington’s diseases template which represent summary about genetics and biomarkers. See links and references in Supplemental materials.

Anatomy biomarkers such as neuron, astrocyte, medium spiny neuron, neuroglia, microglia, and several molecular biomarkers were reported for the HD. However, most of supporting references were conducted on animal models, since it is not possible to research biomarkers in brain tissues in humans. Neurofilament light protein (NEFL) may be promising blood biomarker, because the level of this protein was linked to the severity in Huntington’s disease model (Soylu-Kucharz et al., 2017).

Cellular and molecular mechanisms of disease can be shown as signaling pathways – defects in cascades of transferring the molecular signals in cell. There are 8 pathways presented in the template that were manually built in Pathway Studio software using published studies. Each pathway includes description, links to published articles, and additional information about members (proteins, drugs, cells, etc.).

The central pathway – Huntingtin Gene Mutation in Striatal Neuron – is the disease model (Figure 6) which describes how the mutation in one gene may trigger changing in the cell fate and be reason of the disease. Normal huntingtin (HTT) was shown to play a role in fast axonal transport. Mutant HTT (mHTT), for example, can disrupt binding of the HAP1/DCTN1 protein complex to microtubules and motor complexes and thereby can depress axonal transport and release of Brain-derived neurotrophic factor (BDNF). mHTT binds more strongly to beta-tubulin (microtubules) than does wild-type HTT, thereby accumulating over time on the microtubules. This causes a physical block to transport and gradually makes intracellular transport less efficient.

**Figure 6.**
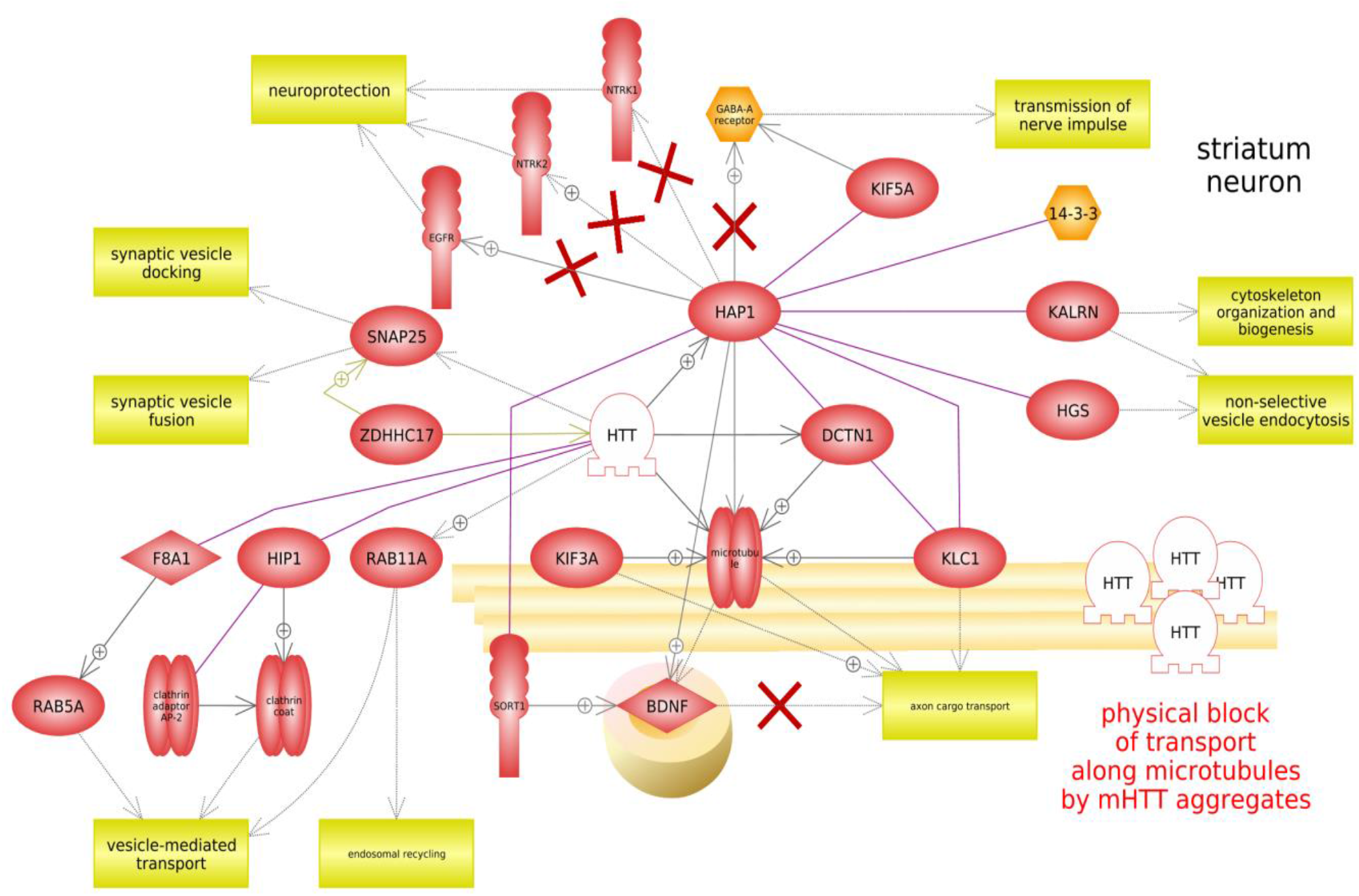
Axonal transport is impaired in neurons due to mutation in HTT. **Signaling description:** Normal huntingtin (HTT) was shown to play a role in fast axonal transport. Mutant HTT (mHTT) can disrupt and gradually makes intracellular transport in neurons less efficient. HAP1, a HTT interacting protein, facilitates the interaction of HTT with dynactin 1 (p150Glued, DCTN1), an accessory partner of dynein/dynactin motor protein complex. HTT/DCTN1/HAP1 protein complex interacts with microtubules to facilitate transport of the vesicle along the axon. It appears that HAP1 acts as a BDNF cargo-carrying molecule. Pro-BDNF forms a complex with HAP1 and sortilin that modulates pro-BDNF trafficking, degradation, and processing. Reduced association among pro-BDNF, HAP1 and SORT1 is observed in the Huntington disease (HD), which may result in impaired BDNF transport. HAP1 is involved in the internalization and recycling of the GABAA receptor. Delivery of GABAARs to synapses is mediated by HAP1-KIF5 and disrupted by mHTT. HAP1 regulates trafficking is of membrane receptors NTRK1, NTRK2 and EGFR, which can be attenuated by mHTT.HTT function in vesicle trafficking is also mediated by palmitoylation. Palmitoylation of HTT by HIP14 (ZDHHC17) is essential for its trafficking and function; it influences HTT localization and protects it from aggregation. Palmitoylation of SNAP25 (a protein, taking part in vesicle fusion and docking) by ZDHHC17 is potentiated in the presence of wild-type HTT. mHTT has prominently reduced affinity to ZDHHC17 that results in a marked reduction in palmitoylation. mHTT also impairs vesicle-mediated transport via interaction with RAB11A, HIP1 and HAP40 (F8A1). 14-3-3 protein interacts with HAP1 and regulates its trafficking. Duo (KALRN) binds to HAP1-binding proteins and regulates cytoskeleton (actin) organization and endocytosis. HAP1 also interacts with hepatocyte growth factor-regulated tyrosine kinase substrate (HRS), which plays a role in the regulation of vesicular trafficking and signal transduction. Mutated genes are shown in white-out style.

**Figure 7.**
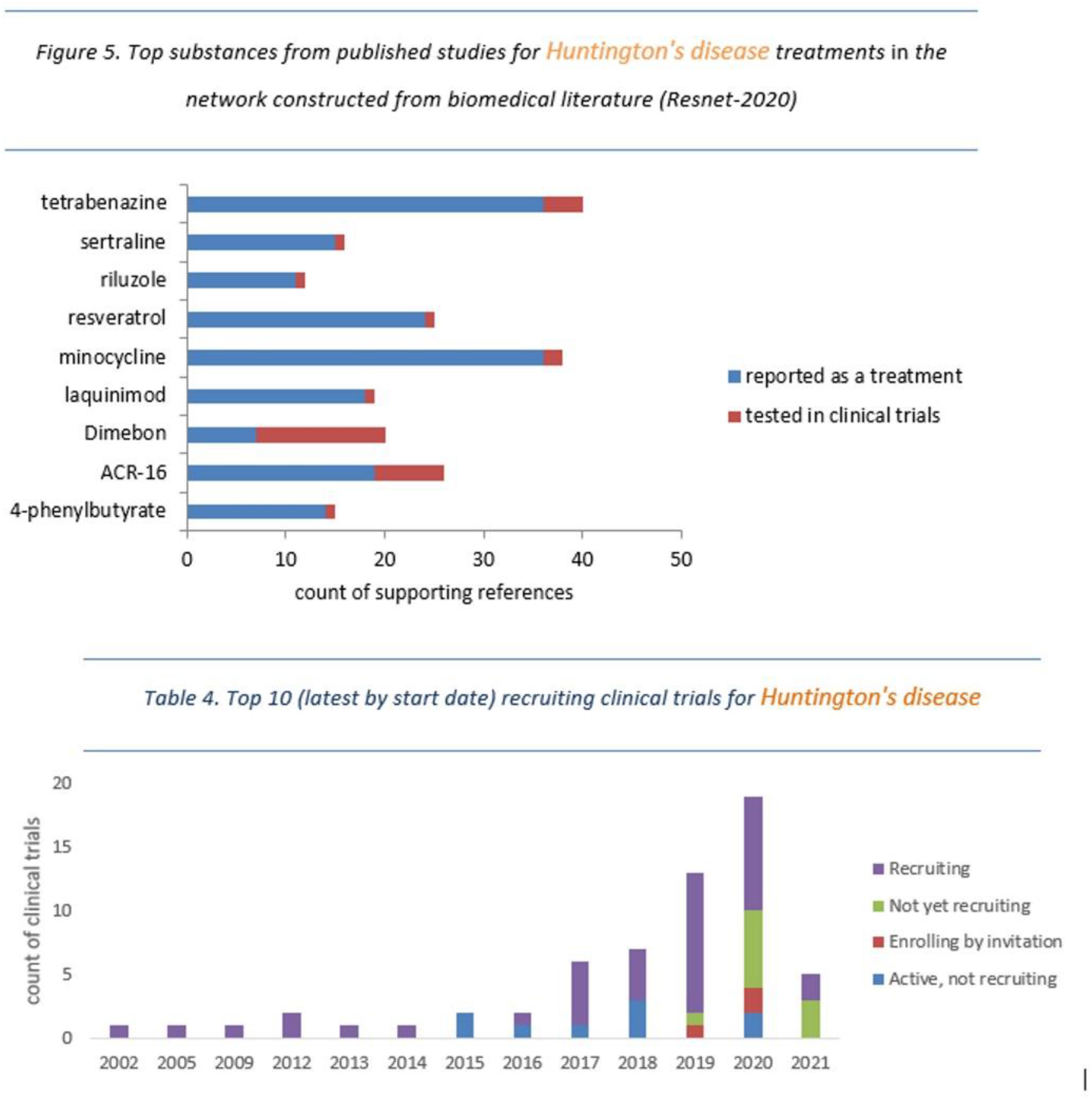
Infographics from Huntington’s diseases template represent the summary of Treatment section. See links and references in Supplemental materials.

Treatment section of the template highlights multidisciplinary approach which should be based on treating symptoms with a view to improving quality of patients’ life because we do not know yet how to eliminate the toxic action of mutant HTT gene on neurons. Multidisciplinary rehabilitation, involving exercise and cognitive training, enhances brain volume in the striatum and prefrontal cortex and improves cognition and motor function (Bartlett et al., 2020). Chorea is treated with dopamine receptor blocking or depleting agents.

Cell replacement therapy (CRT) and gene therapy may be promising ways to find a cure for HD. (Adil et al., 2018; Evers et al., 2018).

Two drugs (tetrabenazine, deutetrabenazine) were found as FDA approved frugs for the treatment of HD with acceptable safety protocols. However, there are many more research and clinical trials for other pharmacological treatments. For example, Dimebon and Pridopidine were actively tested in clinical trials for HD patients.

Last sections of the template included information about the prognosis and cases. Disease progression leads to the complete dependency in daily life; however, heart disease and pneumonia are major risk factors for mortality in patients with Huntington disease. Depression and aggression are also common and need to be treated. However, multiple publications about case studies and experimental treatment options indicate that patients with HD may live long life with right diagnosis and treatment of accompanying illnesses.

### 3.5 RARE MICRODELETION SYNDROMES AND OTHER DISEASE TEMPLATES

Rare microdeletion syndromes (108 rare syndromes from Orphanet and text mining) is a disease group which is easy to detect with karyotype analysis but which is not well covered in the literature and databases (Watson et al., 2014; Weise et al., 2012). We tried to automate the search of information for microdeletion syndromes. Smith-Magenis syndrome, 22q11.2 deletion syndrome, 20p13 microdeletion, 15q11.2 microdeletion syndrome, 14q11.2 microdeletion syndrome had the richest data, however there were not enough information to generate disease templates. Automatically retrieved references and network data for rare microdeletion syndromes as well as for additional 48 rare diseases and several cancers can be found in supplemental materials.

## 4 Summary

In modern biomedical research and data analysis, searching the data and merging results of different types of information (molecular, clinical, pharmacological) is a time-consuming task. We attempted to use automated text mining and querying databases to link the data on the origin, pathogenesis, molecular mechanisms, and approaches to therapy to create templated disease descriptions, which authors may then use as a basis for writing a deeper overview or similar content on the disease of interest. We retrieved information for the top 63 socially significant rare monogenic diseases and 108 microdeletion syndromes. Disease templates can be downloaded and include lists of key disease characteristics, genes, biomarkers, pathways, and drug references to literature data as well as links to public databases. Such sets of information and automated workflow may be useful as most relevant disease overviews, as reference sets for machine learning, and as educational materials for trainings in medicine, biology, or pharmacology. Automated templates may be updated regularly without additional efforts and used as example for building similar workflow with other resources and even with other biomedical terms. Use of commercial Elsevier databases and software is the limitation of this work, since of these are not available to all researchers and it is difficult to estimate their quality and completeness.

## 5 Methods

Data analysis and data merging were done by querying SQL Server database.

Mutated genes were selected by searching Resnet network for “Genetic Change” connections between disease and genes. And search “Functional Association” connections between disease and SNVs from ClinVar database. To search drugs that were reported to tested with the disease in Resnet, we search for “Clinical Trials” and “Regulation” types of connections between disease and drug. Biomarkers were found by searching for proteins, cells, tissues, and organs that connected with disease by “Biomarker” and “Quantitative Change” type of relations. Filters such as number of references and effects of relations were applied.

Top genes and biomarkers for sub-sections can be extracted automatically with Pathway Studio API for any disease name by request. Example of queries are in Supplemental files.

Pathways and network interactions with genes, drugs and biomarkers were built in Pathway Studio 9.0 (desktop version) and are available at https://mammalcedfx.pathwaystudio.com/ and the links in Supplemental files.

Extracting of sentences and citation was done with API of Elsevier Text Mining and using of Advanced Query Language (AQL). Elsevier Text Mining uses Natural Language Processing (NLP) technology that identifies, extracts, indexes, and searches relations in all sentences of indexed documents. ETM has semantic relations-based search. Relations are defined by Subject-Verb-Object triplets. Extracted triplets are normalized to account for various grammatical constructs, including word order, tense, and passive/active voice. AQL allows a user to build a complex query based on different scope operators to look for specific taxonomy and/or free-text terms in particular document sections (title, abstract, text body), co-occurrence levels (semantic relation, sentence, paragraph, document), or as semantic types (subject, verb, object).

Following AQL queries were used:

rel(hemophilia AND prevalence AND Measurement Values); rel({hemophilia} AND obj({birth prevalence} OR {incidence}) AND {Measurement Values}); rel({hemophilia} AND obj({birth prevalence} OR {incidence})); rel({hemophilia} AND “risk factors”); parag(rel({hemophilia} AND “first symptom”) OR rel({hemophilia} AND “early symptoms”)); rel({hemophilia A} AND “major symptoms”); sent(rel({hemophilia}/syn/noexp AND {complications}) AND NOT “treatment”); rel({hemophilia} AND

{mortality}); rel({hemophilia A} AND {differential diagnosis}); rel({hemophilia} AND {diagnostic procedure}); rel({hemophilia} AND “known causes”); rel({hemophilia} AND {therapy} AND NOT {drug therapy}); rel({hemophilia} AND {prognosis}); rel({hemophilia} AND {case report}) OR rel({hemophilia} AND {case study}) OR rel({hemophilia}).

### 5.1 SUPPLEMENTAL MATERIALS

Supplemental materials can be downloaded from ResearchGate (DOI:10.13140/RG.2.2.10990.48960).

The file “Rare Monogenic Diseases.xslx” includes results of ranking rare diseases based on exported data from databases Orphanet and Resnet – 2020.

Rare disease templates can be found in Supplemental materials and also published in Mendeley Data (https://data.mendeley.com) by searching “Elsevier Author Aid Human Diseases Collection”, part 2 and part 3) (Nesterova, 2021).

Author Aid Human Diseases Collection was prepared in 2021 in Elsevier Mendeley Data platform and contains curated materials for 63 rare monogenic diseases, 108 microdeletions, and 7 cancers, as well as for migraine and COVID19. The project is also documented in ResearchGate (https://www.researchgate.net/project/Author-Aid-Human-Diseases-Collection).

Pathways with mechanisms of rare monogenic diseases linked to the top mutated genes and biomarkers and supporting references can be found in Mendeley Data or in Supplemental materials by the name of a disease in files “Pathway Studio exported data”. In addition, links to pathways and their taxonomy can be browsed in Biological Pathway Taxonomy (BPT) (https://bioportal.bioontology.org/ontologies/BPT).

The files with models and pathways in original RNEF format as well as in popular OWL and SBGN formats with all entities and metadata are available by request.

Finally, examples of queries and python code to retrieve network information from ETM can be found on GitHub (https://github.com/nesterova-anastasia/authoraid-diseases).

## Data Availability

Data stored as supplemental files in ResearchGate (DOI:10.13140/RG.2.2.10990.48960).

DOI:10.13140/RG.2.2.10990.48960

